# Understanding The Impact, Reach and Implementation of a Health Systems Intervention to Improve Diabetes and Hypertension Care in Pluralistic Urban Public Primary Care in Bangladesh: A Study Protocol

**DOI:** 10.1101/2025.04.28.25326546

**Authors:** Deepa Barua, Joseph Hicks, Helen Elsey, Mahua Das, Umme Salma Anne, Nabila Binth Jahan, Khaleda Islam, Rahat Chowdhury, Mahmuda Ali, Bassey Ebenso, Tim Ensor, Rumana Huque

**Author notes:** Corresponding Author: Deepa Barua Postal Address: ARK Foundation, Suite C3 and C4, House # 6, Road # 109, Gulshan – 2, Dhaka -1212, Bangladesh Email ID.

## Abstract

**Introduction:** In Bangladesh government provision of primary care in rural areas has seen the development of services for non-communicable diseases, particularly hypertension and diabetes (given their substantial rise in recent decades). However, in the context of cities, which are characterised by a plurality of providers that have sprung up to meet the demands of a rapidly growing urban population, such provision is very limited.

**Methods and analysis:** We will conduct a mixed-methods study, based on the RE-AIM framework, to understand the 1) reach, 2) effectiveness, 3) adoption, 4) implementation, and 5) maintenance (the five RE-AIM domains) of a health systems intervention to strengthen management processes for hypertension and diabetes within government and non-governmental (NGO-run) primary care facilities. To evaluate the effectiveness of the intervention we will use a quasi-experimental, difference-in-differences design. We will recruit 20 purposively selected urban government-run and NGO-run primary care facilities across Dhaka North and South City Corporation areas. Ten facilities will be purposively allocated to an intervention group and receive training and guidance materials on diabetes and hypertension care, based on the WHO Package of Essential Noncommunicable (PEN) Disease Intervention for Primary Care, and the use of an e-health application for patient records. The remaining facilities will be allocated to the existing care group and receive no intervention inputs, with identical data collection processes carried out in both groups. We aim to collect data on 50 patients visiting each facility during a baseline period and at 6- and 12-months after implementing the intervention. Using a standard linear-regression based approach, we will estimate the difference-in-differences effect of the intervention on a primary outcome measuring how many of eight key management processes were appropriately carried out for each patient visiting the facility at 6- and 12-months after implementing the intervention (with their appropriateness based on what should occur according to the intervention guidelines). We will also analyse the effect of the intervention on each management process separately as secondary outcomes. Alongside this design we will collect a range of additional quantitative and qualitative data to evaluate the other RE-AIM domains, using sequential mixed methods approaches, focusing on understanding potential facilitators and barriers in relation to these domains.

**Ethics and Dissemination:** Ethics approval has been received from the Research Governance Committee at the University of Leeds, UK (MREC 21-008) and from the Bangladesh Medical Research Council (BMRCAIREC/20 I 9 -2022 / 485). We will use a variety of channels to share our findings with policy makers, service providers, academicians, and relevant stakeholders.

**Strengths and Limitations of the Study:** *Strengths:*

- This will be the first study to use implementation research methods to understand the delivery of diabetes and hypertension management in the context of the pluralistic urban primary care services of Bangladesh.
- The use of the RE-AIM framework to structure and plan evaluation will help assess multiple dimensions of implementation under real-world conditions therefore providing detailed insights for decision-making on the potential future scale-up of the National Protocol and Simple App within PHC facilities in urban Bangladesh.

*Limitations:*

- Conducting a randomised trial design for effectiveness evaluation will not be possible due to lack of resources
- Planning an evaluation for a longer time scale will be difficult due to project funding timelines

## INTRODUCTION

The World Health Organization (WHO) recommends strengthening primary health care (PHC) systems in all countries to provide fair and efficient care, to contribute to better health outcomes, lower healthcare costs, and to reduce health disparities (1,2). In response to the rising burden of non-communicable diseases (NCDs), the WHO’s *Global Action Plan for the Control of Noncommunicable Diseases 2013–2020* recommended that countries strengthen their health systems and address NCDs through people-centred PHC and universal health coverage (UHC) (3). More specifically, the WHO’s Package of Essential Noncommunicable Disease Interventions for Primary Health Care (4), (from here on referred to as PEN) has been recommended as a ‘best buy’ for low- and middle-income countries (LMICs) seeking to improve the coverage of appropriate services for people with NCDs in primary care settings. PEN includes cost-effective interventions for the early detection and management of NCDs, and has been successfully implemented in rural settings in many LMICs (5).

However, in many LMICs there are significant differences in the health care system between rural and urban areas, often due to national context, government and donor priorities. This is the case in Bangladesh, where this study is located, with the urban PHC system having a different structure and service delivery modality than in rural areas. Specifically, in rural areas PHC service provision is managed solely by the Ministry of Health and Family Welfare (MoHFW). However, while the MoHFW also manages some government-run PHC facilities in urban areas, the Ministry of Local Government, Rural Development and Cooperatives (MoLGRD&C) are responsible by law for the provision of PHC to the urban population, and this is done primarily through contracted NGOs(6).

In rural Bangladesh within each subdistrict a public PHC facility, known as an Upazila Health Complex, provides specialist NCD services, including for hypertension, diabetes and chronic obstructive pulmonary disease, in areas of the facility known as an NCD corner (7). NCD corners operate according to Bangladesh’s *National Protocol for Management of Diabetes and Hypertension* (8) (referred to from here on as the National Protocol), which has been adapted from WHO’s PEN (4) by the MoHFW and guides health workforce on the management of hypertension and diabetes mellitus (from here on referred to as diabetes) at PHC settings (9). However, a recent needs assessment found that urban PHC facilities in Bangladesh lack any equivalent NCD service provision(10–12).

Similarly, given that both hypertension and diabetes are long-term conditions requiring considerable patient follow-up and complex management, a robust individual, patient-level recording system is required and recommended by PEN. Rural PHCs have an NCD patient record-keeping system, either paper-based or digital, although it is not always very effective (9,13), and a digital health information system, named the *Simple App*, is currently being used at some rural PHC facilities with plans for wider roll-out under the supervision of MoHFW. The app is linked to the National Health Dashboard and can thus be used by public health decision-makers to understand the distribution of these conditions and plan programmes and policy in response (13). However, while in urban areas government and NGO PHC facilities have paper-based patient recording systems they are not adapted for NCD patient management (10–12).

As with other LMICs, the implementation of PEN within PHC facilities in urban areas in Bangladesh has not yet occurred, due in part to the complexity of primary care provision in urban areas. However, it is well known that the prevalence of NCDs, particularly hypertension and diabetes, along with their known and potential causes, have risen greatly in LMICs, including Bangladesh, in recent decades, particularly in urban areas (14,15). Therefore, this study aims to address the gap in the evidence base of how to implement PEN within the pluralistic PHC context found in urban Bangladesh, and to inform such work in LMIC urban areas more widely. More specifically, this study aims to evaluate the implementation and effectiveness of an intervention package for urban PHC facilities, both government and NGO run, in the capital city Dhaka, comprising the National Protocol (based on PEN) and the Simple App, which seeks to improve the management of hypertension and diabetes.

Understanding how to implement such an intervention within the complexities of the urban health system in this rapidly expanding and dynamic urban environment requires a multi-dimensional exploration of the implementation process as well as the intervention’s potential effectiveness. Therefore, we will use the RE-AIM framework (16) to structure our evaluation. The RE-AIM framework addresses five broad outcomes relating to the reach, effectiveness, adoption, implementation and maintenance of an intervention. Based on this framework the primary objectives of this study are to:

1. Assess the reach of the intervention to all patients aged ≥40.
2. Assess the effectiveness of the intervention at improving the appropriate management of patients aged ≥40 (according to the National Protocol guidelines).
3. Assess the extent to which the intervention is adopted, and any facilitators or barriers to the uptake of all components of the intervention, by facilities and healthcare providers.
4. Identify the facilitators and barriers to the implementation of the intervention within routine practice, including fidelity to all components of the intervention and any unintended consequences.
5. Assess the extent to which the intervention is maintained, and how successfully, six months after the implementation period, and explore stakeholder perspectives on scale-up of the intervention across urban Bangladesh.

## METHODS AND ANALYSIS

### Study design

We will evaluate the effectiveness of the intervention at improving the appropriate management of patients in relation to hypertension and diabetes, in comparison to existing practice, using a quasi-experimental difference-in-difference design (DiD) (17). This DiD design will collect cross-sectional, patient-level data during three periods: at baseline (one-month prior to implementing the intervention) and then 6-months and 12-months after intervention implementation. The aim is to estimate the intervention’s short and longer-term causal effect on our outcomes, with the 12-month data collection period allowing us to explore the sustainability and maintenance of any intervention effects.

Alongside the DiD study we will evaluate the other domains of the RE-AIM framework, namely the reach, adoption, implementation and maintenance, via a range of quantitative and qualitative data that we will either collect during one or more of the above data collection periods or separately. Below we describe the methodology we will use to evaluate the effectiveness of the intervention, and then we describe how we will evaluate the remaining RE-AIM domains.

### Study setting and study facilities

The study will be conducted in Dhaka, Bangladesh. Bangladesh is a South Asian country, sharing its borders with India and Myanmar, and is divided into eight administrative divisions, including the Dhaka division, which is further divided into 17 districts. One of these districts is Dhaka District, and within Dhaka District the most urbanised area is split into 5 municipalities and 2 city corporations, namely Dhaka North City Corporation (DNCC) and Dhaka South City Corporation (DSCC). This study will be conducted within these two city corporations, which have been chosen due to their high population and densely crowded neighbourhoods, similar to the other 10 city corporations in the country (18).

Within these two city corporations’ residents seeking low-cost or free PHC have two main options, as of 2025. First, they can use one of 19 government-run urban dispensaries (UDs), which provide basic PHC services that are free to the user (see table 1 for their key characteristics). These facilities are managed by the MoHFW. Second, they can use one of 65 NGO clinics, of which 55 provide general PHC services and 10 provide maternal and neonatal focused primary care services, where the user must pay services are subsidised and provided at low cost (see table 1 for their key characteristics). These facilities operate under the MoLGRD&C through the Urban Primary Health Care Services Delivery Project (UPHCSDP). Additionally, residents may also access PHC services from the out-patient departments of tertiary hospitals, but this is only common among residents living near to such facilities. Therefore, in this study we will just be working within UDs and NGO clinics.

**Table 1.** Characteristics of urban dispensaries and NGO clinics.

| Table 1. Characteristics of urban dispensaries and NGO clinics |  |  |
| --- | --- | --- |
| Characteristics | Urban dispensaries | NGO clinics |
| Leadership and governance | • Public/government owned.<br>• Managed and supervised by Dhaka Civil Surgeon Office and Upazila Health Care (within the Dhaka Civil Surgeon Office), under the MoHFW.<br>• The clinic in-charge is a medical doctor and is responsible for managing the clinic in addition to providing clinical care. | • Owned by national NGOs and part of a public-private partnership.<br>• Managed and supervised by the Project Director Office of Urban Primary Health Care Service Delivery Project, under the DNCC, and by the DSCC, under the MoLGRD&C, and by the NGOs.<br>• The clinic in-charge is a medical doctor and is responsible for managing the clinic in addition to providing clinical care. |
| Service delivery | • Services focus on all health issues, especially communicable diseases.<br>• Diagnostic or lab equipment are not available other than a blood pressure machine and a weight machine. | • Services focus on maternity services and vaccination.<br>• Wide range of diagnostic services are typically available including for NCDs, such as hypertension and diabetes, plus blood testing, ultrasound services etc. |
| Health system financing | • All services are free of cost.<br>• Funded by MoHFW.<br>• No bidding process. | • Ultra poor patients attending these facilities are identified through a survey conducted under the MoLGRD. These patients are then provided with an essential health card, commonly known as a red card, through which they can receive all services and medicines for free. Other patients who do not have the |
|  |  | card can obtain subsidised services. Furthermore, the medical officers at these clinics can provide a discount up to 15% on services and medicines as per their judgement.<br>• Funded through a soft loan granted by the Asian Development Bank, which is managed by the MoLGRD&C.<br>• NGOs are selected through a bidding process. |
| Health workforce | • 4 to 6 staff in each clinic, typically: 2 doctors, 2 sub-assistant community medical officers (medical assistants), 1 pharmacist, and 1 maintenance staff. | • 12 to 14 staff in each clinic, typically: 1 doctor, 3 paramedics, 1 counsellor, 1 administrator, 1 management information specialist, 1 receptionist, 1 lab technician, 4 service promoters, and 1 field supervisor. |
| Medications | • Hypertension and/or diabetes medication may or may not be available, but if available it is unlikely to be as per the National Protocol guidelines. | • Some pregnancy-specific hypertension medication is available but not hypertension medication recommended by the National Protocol, and no diabetes medication is available. |
| Health information systems | • Paper-based patient record system exists but is not adapted for managing NCDs such as hypertension and diabetes.<br>• Provide monthly service provision reports to the Dhaka Civil Surgeon Office. | • Paper-based patient record system exists but is not adapted for managing NCDs such as hypertension and diabetes.<br>• Provide quarterly service provision reports to the Project Director Office. |
|  | • Patient feedback system is not available. | • Patient feedback system is available. |
| DNCC = Dhaka North City Corporation. DSCC = Dhaka South City Corporation. MoHFW = Ministry of Health and Family Welfare. MoLGRD&C = Ministry of Local Government, Rural Development and Cooperatives. |  |  |

The UPHCSDP programme has been delivered through four phases, with the aim of supporting Bangladesh’s urban primary health system by increasing the accessibility of PHC to the urban poor. The programme has been found to be effective in improving child and maternal health indicators, through increased antenatal care coverage, skilled birth attendance, and breast-feeding practices, and to reduce diarrhoea and acute respiratory infection prevalence in children. Additionally, the programme has also been shown to be effective at decreasing sexually transmitted infections through improved HIV/AIDS awareness and increased prevalence of contraceptive use (19).

However, a recent needs assessment conducted by our team assessed the preparedness of UDs and NGO clinics to deliver hypertension and diabetes management services (10–12). In general, the level of preparedness and ability to deliver hypertension and diabetes management services was found to be low. A more detailed summary of these facilities’ characteristics, with a focus on the hypertension and diabetes services, are given in table 1 below, with further discussion of the needs assessment in the *Intervention* section below.

### Target population

Our target population are individuals aged 40 and above who utilise UDs and/or NGO clinics present in urban areas of Bangladesh, and the UDs, NGO clinics, and the medical staff of those facilities who are involved in managing patients in relation to hypertension and diabetes.

### Study facility eligibility criteria

We have minimal eligibility criteria for study facilities to maximise the generalisability of our findings.

Our eligibility criteria for UDs and NGO clinics are:

- Facilities must have been operating consistently for at least a year.
- Facilities must have been providing health care services to all types of patients and not restricted to subgroups such as children, pregnant women and newborn babies.
- Facilities present in general population areas and not inside government-restricted working areas.
- Facilities must have a typical patient load of at least 50 eligible patients per day.
- Facilities must have at least one person in each of the following health workforce roles:

º Medical officer / doctor.
º Medical assistant (for UDs this would be a Sub-Assistant Community Medical Officer [commonly known as a SACMO], and for NGO clinics this would be a paramedic).
º Pharmacist.
- Facilities with at least 3 rooms.
- Facilities not existing within 3 km of each other.

### Patient eligibility criteria

We also only have one eligibility criteria to maximise the generalisability of our findings: patients must be aged 40 or above. This age limit comes from the National Protocol guidance that all patients aged 40 or above should be screened for hypertension and diabetes.

### Intervention description

#### Context of the intervention package

Prior to the development of the intervention package, as previously discussed, we carried out a needs assessment to identify the existing gaps within hypertension and diabetes management in the urban PHC system using the WHO Health Systems Building Blocks framework (11,12). In this assessment, we identified gaps across all building blocks. Specifically, within urban UD and NGO facilities there was often a lack of clinical training and guidelines on hypertension and diabetes management, a lack of screening for hypertension and diabetes, a lack of equipment to diagnose diabetes, a lack of counselling on how to prevent and/or manage hypertension and diabetes, and a lack of follow-up and referral processes for patient management.

The findings of the assessment were then presented to relevant stakeholders, specifically representatives from the NCDC Programme, the MOHFW, the Urban PHC Management Unit under the MoLGRD&C, urban health experts and academics. During this meeting it was agreed with the Government stakeholders that we would prioritise addressing gaps in service delivery, the health workforce and health information systems. The intervention package was then decided upon collaboratively with the government stakeholders as a potential solution to the identified key gaps. It was decided that the intervention would include three main components.

1. Provision of the National Protocol guidelines on managing hypertension and diabetes to clinical staff at urban PHC facilities.
2. Introduction of a digital health information system to manage patients with hypertension and diabetes (i.e. diagnosis, prescription, follow-up and referral recoding) at urban PHC facilities.
3. Training relevant staff at urban PHC facilities on how to manage hypertension and diabetes using the National Protocol guidelines and on how to use the patient health information system.

#### Intervention component details

##### 1. National Protocol for Management of High Blood Pressure and Diabetes

### As previously mentioned, the National Protocol has been adapted from the WHO’s PEN

(8). The National Protocol has been introduced by the MoHFW in Bangladesh, and is currently being implemented across rural PHC facilities (9). The National Protocol provides clinical guidelines for the screening and assessment of all those aged 40 and above to identify those at risk of developing NCDs, specifically cardiovascular diseases, hypertension and diabetes. It also provides clinical guidelines regarding the management of these within PHC settings. Through set diagnostic criteria for hypertension and diabetes, it states the 1^st^, 2^nd^ and 3^rd^ line of treatment medications, which are low cost and usually easily accessible. It also provides guidance on when patients should be followed up, depending on the hypertensive and diabetic status of the patient, or referred, in the case of serious/complicated cases. The National Protocol also provides clinical guidance on brief counselling that should be provided to patients regarding improved lifestyle behaviours, particularly in relation to diet, exercise, and the use of tobacco and alcohol, to prevent and/or manage cardiovascular disease, hypertension and diabetes. This counselling is designed to be delivered within short consultations that are deemed feasible within the often-heavy patient loads in urban PHC facilities.

#### 2. Patient health information system: Simple App

The international NGO *Resolve to Save Lives* developed and introduced a digital health information system, named Simple App, to several LMICs, including Bangladesh, in association with the *National Heart Foundation* (13). The app has been developed for hypertension and diabetes programmes with the goal of supporting patients to keep their blood pressure and blood glucose levels under control by recording individual patient data, (see table 2 below for an overview of the data collected). The app is currently being used at rural PHC facilities in 25 districts under the supervision of MoHFW and is linked to the National Health Dashboard (13) but has yet to be introduced to urban PHC facilities. The app runs on the Android operating system and so can be used on many smartphones and tablet computers.

**Table 2.** Data recorded by Simple App.

| Table 2. Data recorded by Simple App |  |
| --- | --- |
| Socio demographic factors | Age |
|  | Sex |
|  | Address |
| Clinical data | Medical history of whether the patient has suffered from: |
|  | Heart Attack |
|  | Stroke |
|  | Kidney disease |
|  | Blood Pressure measurement |
|  | Blood Glucose measurement |
|  | Hypertension / Diabetes mellitus diagnosis |
| Treatment | Drugs prescribed for hypertension / diabetes mellitus |
| Advice | Next follow up date |
|  | Referral status |

#### 3. Training on the National Protocol and Simple App

A four-day training programme will be provided to a doctor and a medical assistant from each urban PHC facility in the intervention group, with the individuals selected by their respective management authorities. Trainers will be recommended by the Non-Communicable Disease Control (NCDC) Programme and will comprise diabetes and hypertension specialists, urban health experts, and experts on the use of the Simple App. During the first three days trainees will be trained on the National Protocol, specifically around NCD risk factors, screening, management, lifestyle counselling, follow up and referral. The training will be interactive and include role play scenarios to enable health professionals to practice the management of patients, as per the National Protocol. On the fourth day trainees will be trained on the use of the Simple App for registration and recording of patient with hypertension and diabetes.

See table 3 for a comparison between the intervention components and the corresponding components (or lack of) in the existing care group.

**Table 3.** Intervention components and corresponding existing care comparison within study facilities.

|  |  |  |
| --- | --- | --- |
| Intervention component | Intervention group | Existing care group |
| Hypertension and diabetes management | National Protocol guidelines for managing hypertension and diabetes will be made available at urban PHC facilities. | No guidelines currently available. |
|  | One-page summary of the National Protocol will be made available at urban PHC facilities. | Not currently available. |
| | All patients aged $\geq 40$ attending the facility will be screened by a medical officer/medical assistant for hypertension and diabetes, via a systemic and defined screening process, as per the National Protocol. Those at risk of developing, or who have developed, hypertension and/or diabetes will be identified via this screening. | Screening of patients aged $\geq 40$ for hypertension and diabetes is rarely done/there is no systematic screening process. Consequently, patients at risk of developing, or who have developed, hypertension and/or diabetes are rarely identified. |
| | All patients aged $\geq 40$ will be counselled on lifestyle behaviour modification, as per the National Protocol. | Usually, only patients aged $\geq 40$ who are already diagnosed with hypertension and/or diabetes may be counselled on lifestyle behaviour modification by the medical officer/medical assistant, and this will be as per their initial training, often leading to non-uniformity across facilities. |
| | All patients aged $\geq 40$ will be advised about follow up by the medical officer/medical assistant, as per the National Protocol. | Usually, only patients aged $\geq 40$ who are already diagnosed with hypertension and/or diabetes are advised about follow up by the medical officer/medical assistant, and this will be as per their initial training, often leading to non-uniformity across facilities. |
| | Any patients aged $\geq 40$ who are newly diagnosed as having hypertension and/or diabetes will be prescribed with medications by the medical officer/medical assistant, as per the National | Patients aged $\geq 40$ who are newly diagnosed with hypertension and/or diabetes will be prescribed medications by the medical officer/medical assistant, but |
|  | Protocol. | this will be as per their initial training, often leading to non-uniformity across facilities. |
| | Any patients aged $\geq 40$ who are known to have existing hypertension and/or diabetes and are already on medications and have controlled blood pressure and/or blood glucose levels will be asked to continue their already prescribed medicines by the medical officer/medical assistant. | Patients aged $\geq 40$ who are known to have existing hypertension and/or diabetes will usually be asked to continue taking their previously prescribed medications by the medical officer/medical assistant. |
| Patient health information system for hypertension and diabetes management | All patients aged $\geq 40$ who are newly or previously diagnosed with hypertension and/or diabetes will be recorded in the Simple App by the medical officer/medical assistant. | All patients aged $\geq 40$ who are newly or previously diagnosed with hypertension and/or diabetes will be recorded in the existing paper-based patient records system by the medical officer/medical assistant. |
| Training of the urban PHC workforce to manage hypertension and diabetes | Medical officers and medical assistants will receive a three-day training programme on how to implement and use the National Protocol for managing hypertension and diabetes. | No such training is currently given or will be given as part of this study. |
|  | Medical officers and medical assistants will receive a one-day training session on the use of Simple App. |  |

##### Intervention treatment pathway

Medical assistants (SACMO/paramedics) will screen all patients aged 40 and above, who attend the urban PHC facilities, as per the National Protocol. Based on the results of the screening, a doctor will then identify those at risk of developing cardiovascular disease, hypertension and/or diabetes, or diagnose those who have hypertension and/or diabetes. Patients identified as being at risk of developing one or more of these conditions, or as having one or more of these conditions, will be managed as guided by the National Protocol. All patients will be advised to have a follow-up appointment, with those at risk/diagnose with one or more of these conditions advised to have this sooner. And those patients diagnosed with one or more of these conditions will be prescribed with appropriate medicines (as per the National Protocol) and/or referred elsewhere if needed (in the case of severe cases). All patients will then be sent back to the medical assistant to receive face-to-face lifestyle counselling, based on their NCD status, in line with the National Protocol. Additionally, the medical assistant will register those identified with one or more of these conditions in the Simple App.

See figures 1-3 for an overview of the patient treatment pathways under the intervention.

**Figure 1.**
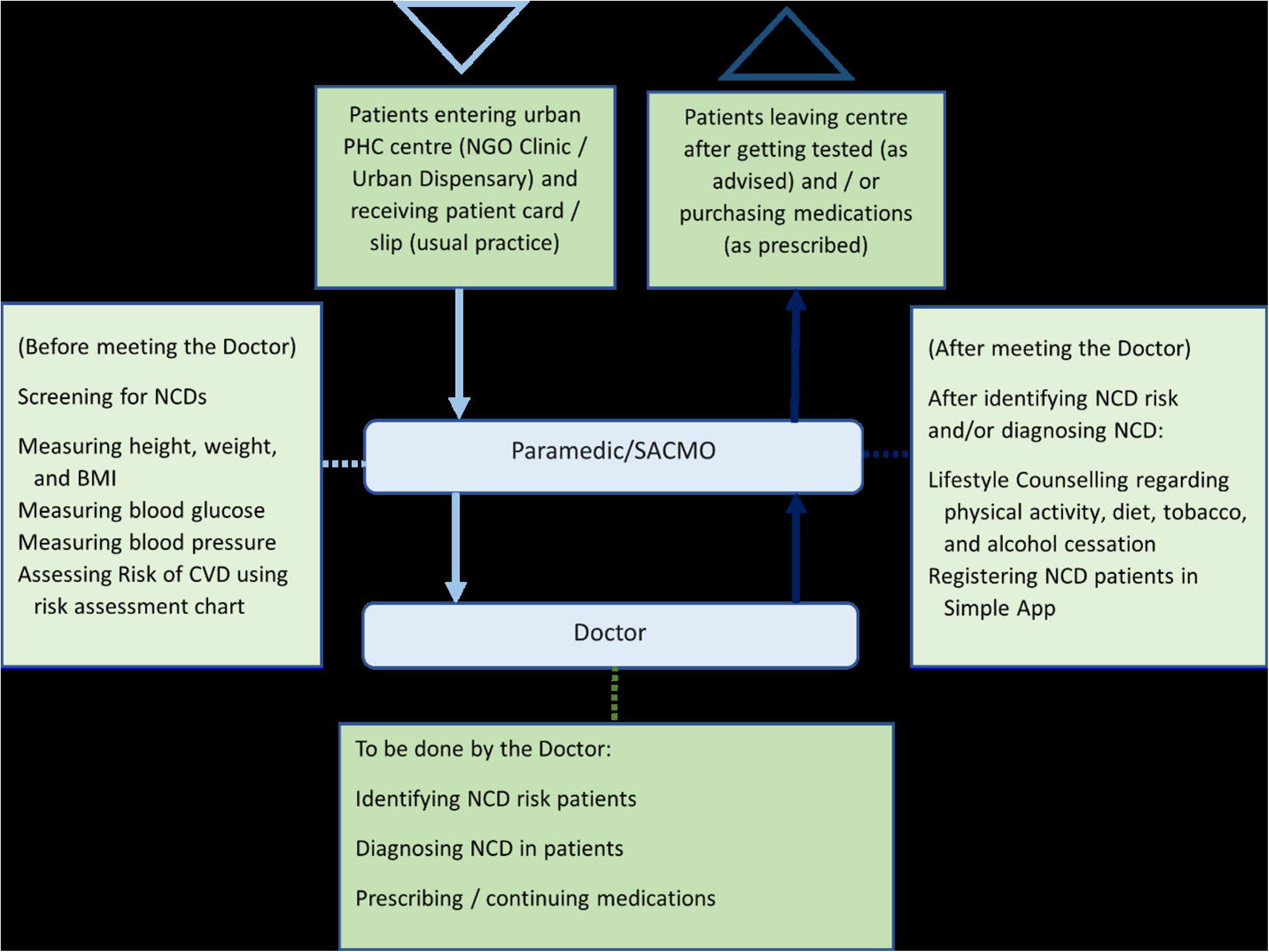
Patient pathway through the intervention.

**Figure 2.**
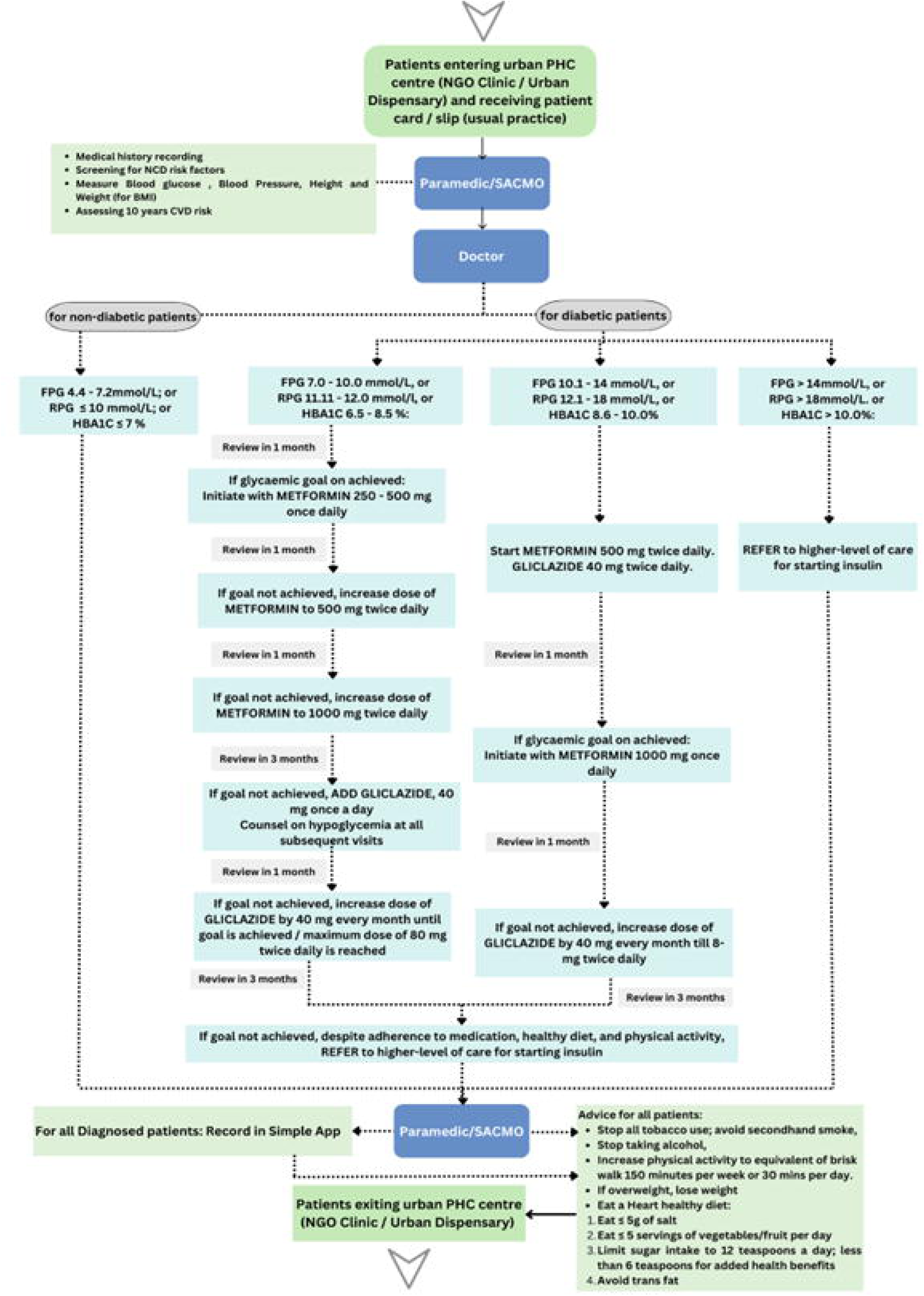
Diabetes management pathway.

**Figure 3.**
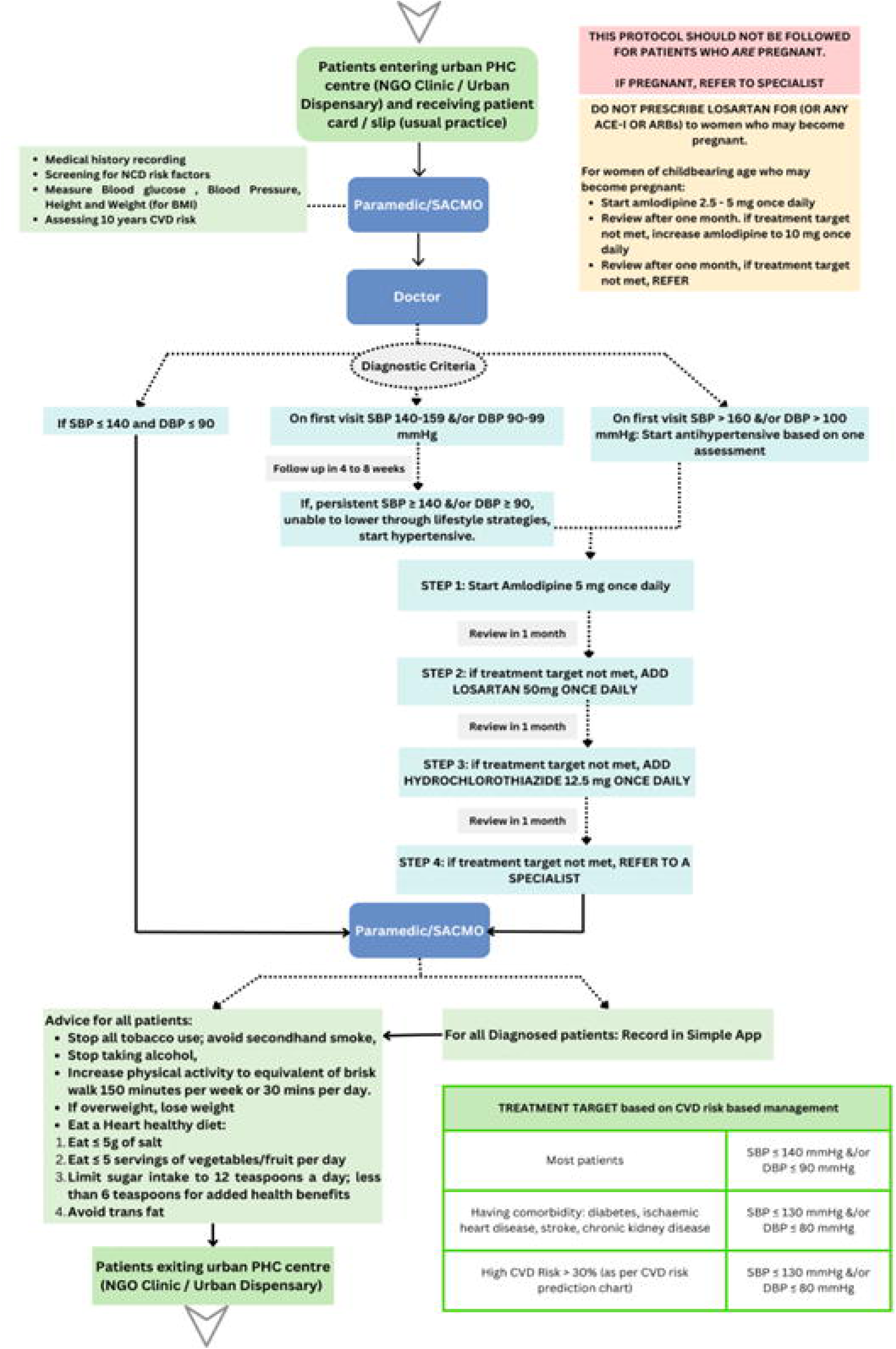
Hypertension management pathway.

## Outcomes

We will collect the following primary and secondary outcomes at the patient-level at baseline and at 6- and 12-months after implementing the intervention, with all outcomes relating to appropriate management of patients in relation to diabetes and hypertension.

### Primary outcome

Our primary outcome will measure the extent to which all patients aged ≥40 attending study facilities (who may or may not be suffering from hypertension and/or diabetes) are appropriately managed as per treatment guidelines in the National Protocol. We will measure this outcome at baseline and at 6-months and 12-months post-intervention. The outcome will allow us to understand whether, and to what extent, the intervention has improved the overall level of appropriate management (as per the National Protocol guidelines) for patients aged ≥40. Although this outcome may be affected by the digital patient health information system component of the overall intervention we do not expect any causal effect of this component of the intervention on this outcome. Therefore, we will assess the direct effects of this component of the intervention separately via a separate outcome, for reasons further explained below.

The primary outcome will look at eight key management processes and whether each one was appropriately carried out or not, according to the National Protocol guidelines. For each patient the outcome will measure how many of these eight management processes were appropriately carried out or not as a 0-8 score, and as described below we will present the results of our analyses of this outcome in terms of the percentage of management processes appropriately carried out. Below we describe these management processes and define how we classify whether the process has been appropriately or not appropriately carried out.

1. Whether the patient has been appropriately screened for hypertension. The patient is considered to have been appropriately screened for hypertension if the patient’s systolic blood pressure (SBP) and diastolic blood pressure (DPB) has been measured, and inappropriately screened if their SBP and/or DBP have not been measured.
2. Whether the patient has been appropriately screened for diabetes. The patient is considered to have been appropriately screened for diabetes if the patient’s fasting plasma glucose (FPG) or random plasma glucose (RPG) has been measured, and inappropriately screened if either their FPG or their RPG have not been measured.
3. Whether the patient has been appropriately diagnosed in relation to their hypertension status. Diagnosis in relation to a patient’s hypertension status will be considered appropriately carried out if one of the below sets of criteria are met for the patient and inappropriately carried out if none of the below sets of criteria are met for the patient.

º The patient i) was not previously diagnosed by a healthcare provider with hypertension, ii) had a SBP <140 mmHg and a DBP <90 mmHg, and iii) was not diagnosed with hypertension.
º The patient i) was previously diagnosed by a healthcare provider with hypertension, ii) was already on antihypertensive medications, and iii) was diagnosed with hypertension.
º The patient i) was previously diagnosed by a healthcare provider with hypertension, ii) was not already on any antihypertensive medications, iii) had a SBP of 140-159 mmHg and/or a DBP 90-100 mmHg, and iv) was diagnosed with hypertension.
º The patient i) was not previously diagnosed by a healthcare provider with diabetes, ii) had a SBP ≥160 mmHg and/or a DBP ≥100 mmHg, and iii) was diagnosed with hypertension.
4. Whether the patient has been appropriately diagnosed in relation to their diabetes status. Diagnosis in relation to a patient’s diabetes status will be considered appropriately carried out if one of the below sets of criteria are met for the patient and inappropriately carried out if none of the below sets of criteria are met for the patient.

º The patient i) was not previously diagnosed by a healthcare provider with diabetes, ii) had a FPG <7 mmol/L or a RPG of <11.1 mmol/L, and iii) was not diagnosed with diabetes.
º The patient i) was previously diagnosed by a healthcare provider with diabetes, ii) was already on antidiabetic medications, and iii) was diagnosed with diabetes.
º The patient i) was previously diagnosed by a healthcare provider with diabetes, ii) was not currently on any antidiabetic medications, iii) had a FPG of 7-10 mmol/L or a RPG of 11.1-12 mmol/L, and iv) was diagnosed with diabetes.
º The patient i) was not previously diagnosed by a healthcare provider with diabetes, ii) had a FPG ≥10.1 mmol/L or a RPG ≥12.1 mmol/L, and iii) was diagnosed with diabetes.
5. Whether the patient has been appropriately prescribed the correct medications, or appropriately not prescribed any medications, in relation to their hypertension status. Prescription in relation to a patient’s hypertension status will be considered appropriately carried out if one of the below sets of criteria are met for the patient and inappropriately carried out if none of the below sets of criteria are met for the patient.
  º The patient i) was not previously on any antihypertensive medications, ii) had a SBP <140 mmHg and a DBP <90 mmHg, and iii) was not prescribed any antihypertensive medications.
  º The patient i) was not previously diagnosed by a healthcare provider with hypertension, ii) had a SBP of 140-159 mmHg and/or a DBP 90-99 mmHg, and iii) was not prescribed any antihypertensive medications.
  º The patient i) was previously on antihypertensive medications, ii) had a SBP <140 mmHg and/or a DBP < 90 mmHg, and iii) was prescribed the antihypertensive medications that they were already on.
  º The patient i) was previously diagnosed by a healthcare provider with hypertension, ii) was not already on any antihypertensive medications, iii) had a SBP of 140-159 mmHg and/or a DBP 90-99 mmHg, and iv) was prescribed with amlodipine.
  º The patient i) was not previously diagnosed by a healthcare provider with hypertension, ii) had a SBP ≥ 160 mmHg and/or DBP ≥ 100 mmHg, and iii) was prescribed with amlodipine.
  º The patient i) was already on amlodipine, and ii) had a SBP >140 mmHg and/or a DBP >90 mmHg, and iii) was prescribed with losartan.
  º The patient i) was already on amlodipine and losartan, and ii) had a SBP >140 and a DBP >90, and iii) was prescribed with hydrochlorothiazide.
6. Whether the patient has been appropriately prescribed the correct medications, or appropriately not prescribed any medications, in relation to their diabetes status. Prescription in relation to a patient’s diabetes status will be considered appropriately carried out if one of the below sets of criteria are met for the patient and inappropriately carried out if none of the below sets of criteria are met for the patient.

º The patient i) was not previously diagnosed by a healthcare provider with diabetes, ii) had a FPG ≤10 mmol/L or a RPG ≤12 mmol/L, and iii) was not prescribed any diabetes medications..
º he patient i) was previously on antidiabetic medications, ii) had a FPG <7 mmol/L or a RPG <11.1 mmol/L, and iii) was prescribed the antidiabetic medications that they were already on.
º The patient i) was previously diagnosed by a healthcare provider with diabetes, ii) was not currently on any antidiabetic medications, iii) had a FPG of 7-10.0 mmol/L or a RPG of 11.1-12.0 mmol/L, and iv) was prescribed with metformin.
º The patient i) was not previously diagnosed by a healthcare provider with diabetes, ii) had a FPG of 10.1-14 mmol/L or a RPG of 12.1-18 mmol/L, and iii) was prescribed with metformin and gliclazide.
º The patient i) was already on metformin, ii) had a FPG >7 mmol/L or a RPG >10 mmol/L, and iii) was prescribed with metformin or metformin and gliclazide.
º The patient i) was already on metformin and gliclazide, ii) had a FPG >7 mmol/L or a RPG >10 mmol/L, and iii) was prescribed with metformin and gliclazide or also with insulin.
º The patient i) had a FPG >14 mmol/L or a RPG >18 mmol/L and ii) was prescribed with insulin.
7. Whether the patient has been appropriately counselled about healthy lifestyle behaviours at least in terms of diet and exercise. Counselling in relation to healthy lifestyle behaviours at least in terms of diet and exercise will be considered appropriately carried out if the patient has been counselled about healthy lifestyle behaviours in terms of diet and exercise, and inappropriately carried out if they have not been counselled about healthy lifestyle behaviours in terms of diet and/or exercise.
8. Whether the patient has been appropriately advised about follow-up given their current hypertension and/or diabetes status and risk. Follow-up will be considered appropriately carried out if one of the below sets of criteria are met for the patient and inappropriately carried out if none of the below sets of criteria are met for the patient.

º The patient i) was not previously diagnosed by a healthcare provider with hypertension and/or diabetes, ii) was not diagnosed with hypertension and/or diabetes, and iii) was advised to return for a follow-up in 6-12 months.
º The patient i) was not previously diagnosed by a healthcare provider with hypertension, ii) had a SBP of 140-159 mmHg and/or a DBP of 90-99, and iii) was advised to return for a follow-up in 1-2 months.
º The patient i) was not previously diagnosed by a healthcare provider with diabetes, ii) had a FPG of 7.0-10.0 mmol/L or a RPG of 11.1-12.0 mmol/L, and iii) was advised to return for a follow-up in 1 month.
º The patient i) was prescribed with antihypertensive medications and ii) was advised to return for a follow-up in 1 month.
º The patient i) was prescribed metformin with a dose up to 500 mg twice daily and ii) was advised to return for a follow-up in 1 month.
º The patient i) was prescribed with metformin with a dose of 1000 mg twice daily and ii) was advised to return for a follow-up in 3 months.
º The patient i) was prescribed with metformin with a dose of 1000 mg twice daily and gliclazide with a dose up to 120 mg and ii) was advised to return for a follow-up in 1 month.
º The patient i) was prescribed with metformin with a dose of 1000 mg twice daily and gliclazide with a dose >120 and ≤160 mg and ii) was advised to return for a follow-up in 3 months.

#### Secondary outcomes

We will also treat each of the eight binary components of the primary outcome score as eight separate secondary outcomes to allow us to explore how each of the different patient management processes targeted by the intervention have or have not changed under the intervention.

#### Referral

We will also collect a binary secondary outcome measuring whether a patient has been appropriately referred, but this will be restricted to those patients who are newly diagnosed with hypertension and/or diabetes or are known to already have either condition. We will measure this outcome at baseline and at 6-months and 12-months post-intervention. Referral will be considered appropriately carried out if one of the below sets of criteria are met for the patient and inappropriately carried out if none of the below sets of criteria are met for the patient.

º The patient i) had a SBP >200 mmHg and/or a DBP >120 mmHg and ii) was referred.
º The patient i) had a SBP >180 mmHg and/or a DBP >110 mmHg, ii) a severe headache, chest pain, shortness of breath, blurred vision, nausea, vomiting, or signs of heart failure, and iii) was referred.
º The patient had a SBP >140 mmHg and/or a DBP >90 mmHg, ii) was already on three antihypertensives and iii) was referred.
º The patient i) had a RPG ≥18 mmol/L and ii) was referred.
º The patient i) had a FPG >7.2 mmol or RPG >10 mmol, ii) was already on the maximum dose of metformin and gliclazide, and iii) was referred.

#### Simple App utilisation

We will also collect a binary outcome measuring whether patients who are newly diagnosed with hypertension and/or diabetes or who are existing cases are registered into the Simple App or not. We will measure this outcome at baseline and at 6-months and 12-months post-intervention. However, this outcome will only be collected for patients in intervention-group facilities, because the Simple App is not being implemented in the existing care group. See the *Statistical analyses* section below for how this outcome will be therefore analysed.

#### Health care provider knowledge

Additionally, we will measure the level of knowledge about the appropriate management of hypertension and diabetes, according to the National Protocol guidelines, among all health care providers (HCPs) from intervention group facilities who receive the intervention training. Their level of knowledge will be measured as the percentage of correctly answered questions assessed via a multiple-choice questionnaire, based on the WHO’s knowledge assessment tool for PEN. The knowledge test will have around 25 questions around case-based scenarios. The questions will be around hypertension and diabetes risk factors, lifestyle modification counselling, medications, follow up and referral. The knowledge test will be given before and at the end of the intervention training programme.

### Sample size

We based our sample size on being able to detect a given difference in our primary outcome between the intervention and comparison groups at 6-months post-intervention. We assumed we would have 4 UDs and 6 NGO clinics per treatment group, which was chosen based on the constraints of our resources and timeframes. We assumed we would be able to recruit 50 patients per facility at baseline and at 6-months post-intervention, based on patient flow data and our resources and timeframes. We assumed the primary outcome of appropriate management would be 12.5% (1/8 management processes appropriately carried out) at baseline across both treatment groups (based on contextual knowledge and our facility needs assessment work) and would remain at 12.5% in the existing care group. We assumed an intra-cluster correlation coefficient of 0.1, based on a conservative rounding up of intra-cluster correlation coefficient estimates (computed via the analysis of variance method) taken from our previous research into Bangladesh’s rural, public, primary care facilities known as community clinics, where we also looked at a similar appropriate management outcome (20).

We then used a simulation-based approach to estimate the power we would expect given these assumptions and different assumptions about the increase in the primary outcome level in the intervention group. For each scenario this involved simulating primary outcome data for 50 patients in each of the 10 facilities in each treatment group for each study period, with a 12.5% outcome level in the existing care group at baseline and 6-months and a 12.5% outcome level in the intervention group at baseline but varying the assumed outcome level in the intervention group at 6-months in each different scenario (i.e. the assumed increase in the outcome due to the intervention). For each scenario we ran 5,000 simulations generating 5,000 datasets.

We then analysed each dataset using the standard difference-in-differences linear regression model(21), with covariates for period, treatment group and their interaction, and computing clustered standard errors with the HC3 correction (22) for the model coefficients. For each scenario we then extracted the two-tailed p-value (based on the t-distribution) for the null hypothesis that the interaction term was 0 from all the models fitted to the 5,000 datasets. Assuming an alpha of 0.05 we then computed the proportion of those p-values that were <0.05, as our empirical measure of power. From this we found that if the above assumptions hold we should be able to detect an increase in the appropriate management outcome to 37.5% (3/8), i.e. a 25 percentage point increase, with 80% or greater power (based on obtaining a two-tailed p-value of 0.05 or lower for the null hypothesis test of the interaction term).

### Facility sampling and recruitment

Within the DNCC and DSCC there are 19 UDs. Of these, 12 are within areas accessible to the general population and will be assessed on the other eligibility criteria. Of the remaining 7 facilities 3 are within government working areas, thereby making them less accessible to public, 3 are school health clinics, and 1 is a maternity clinic, and these will all be excluded from the study. Across the two city corporations there are 65 NGO clinics. Of these, 55 are primary health care facilities and will be considered for inclusion in the study. The remaining 10 facilities are comprehensive reproductive health care centres, which specifically cater to pregnant women and newborn babies and will therefore be excluded from the study.

To determine which facilities will be included in the study we will carry out a needs assessment of the available facilities and, taking the results into consideration, consult with the Dhaka Civil Surgeon Office and both Dhaka City Corporations. As per the sample size calculation, they will select 8 UDs and 12 NGO clinics for inclusion in the study on the basis of the needs assessment, which facilities they have a good relationship with and (following consultation) which facility managers are willing and interested to participate.

### Facility and staff consent

We will verbally brief all health facility managers from the selected PHC facilities on the project and give them an information sheet about what it would involve for them and their facility, staff and patients. For all managers this information will cover data collection methods, while for those managers in the intervention group the information will also cover the intervention package. If the facility managers consent to be a part of the study, they will be asked to sign a consent form. We will also verbally brief health care providers (i.e. the doctor and medical assistant from each facility), responsible for managing patients, on the project, and provide them with an information sheet. For all staff this information will cover data collection methods, while for those staff in the intervention group the information will also cover the intervention package. Those who agree to participate will be asked to sign a consent form.

### Facility allocation

We will ensure that there are an equal number of UDs and NGO clinics per treatment group, as per figure 4. As described above, the Dhaka Civil Surgeon Office and both Dhaka City Corporations will decide which facilities to include, and they will also decide which facilities to allocate to which treatment group based on which facilities they believe will be willing and open to implementing the intervention. Figure 4. Study facility allocation details.

**Figure 4.**
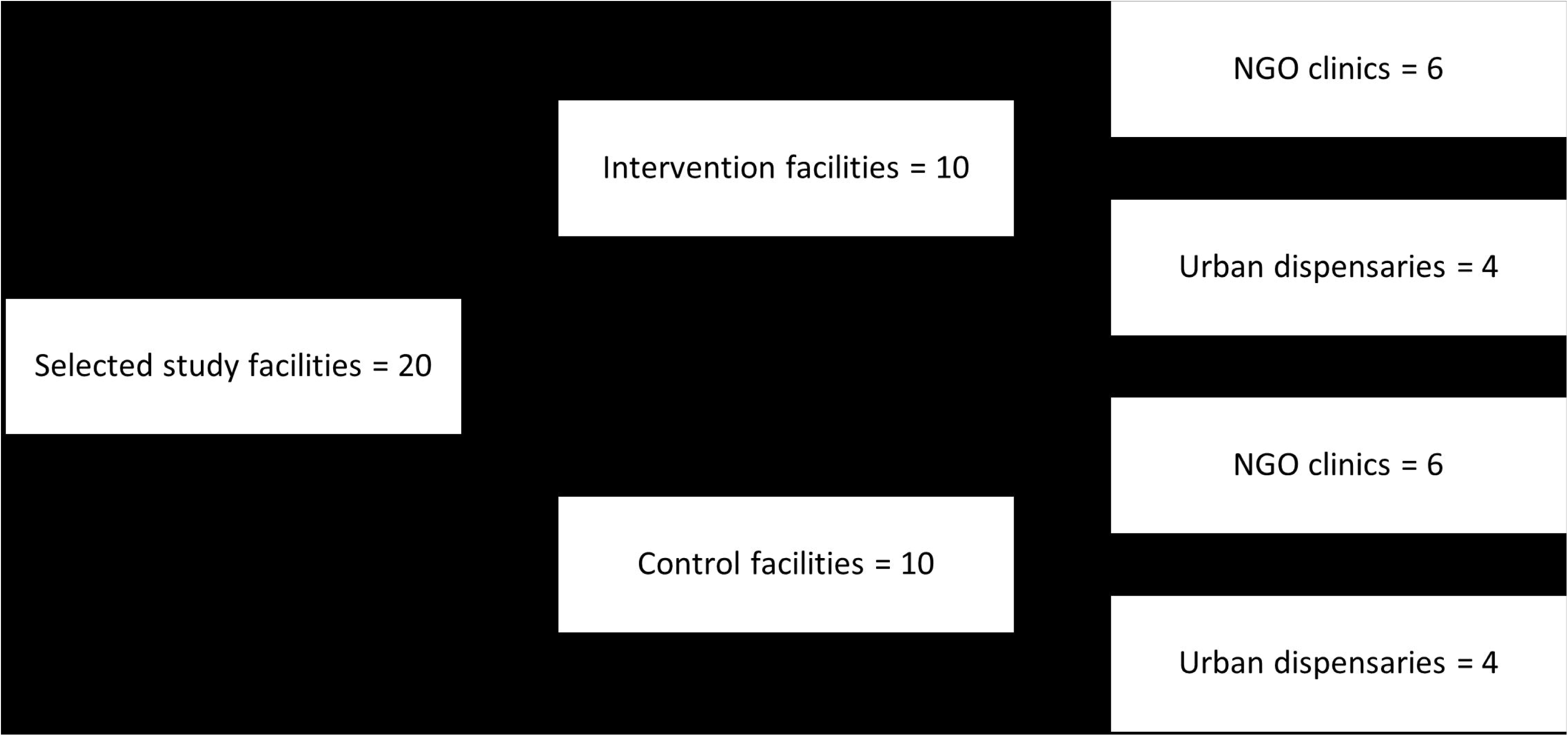
Study facility allocation details.

### Patient sampling and recruitment

We will collect all study data from patients via visits to the study facilities when those patients are attending for treatment. Specifically, during the data collection periods research assistants will be present at the study facilities six days-a-week, from Saturday to Thursday, as per their operating days. One research assistant will be placed at each study facility. Research assistants stationed at the UDs will be there from 8.30 am to 2.20 pm, and those stationed at the NGO clinics will be present from 9.00 am to 4.00 pm, in line with the operating hours of the respective facilities. During each data collection period patients who meet the eligibility criteria and consent to be a part of the study will be consecutively recruited until the desired sample size for that facility is reached.

### Patient consent

During each data collection period, we will invite eligible patients who enter health facilities to participate in the study. Those who are interested will be verbally briefed on the project and provided with an information sheet. If the patient is unable to read the information sheet will be read to them. For all patients this information will cover the data collection methods, while for those patients in the intervention group the information will also cover the intervention package that has been implemented in the facility. If the patient agrees to participate, we will take either written or (if they are illiterate) verbal informed consent to collect data during their consultation and, to access their NCD related records from the patient register, and to access their prescription information (when necessary). Patients will be informed that they can withdraw from the study at any time until their data is analysed. If they withdraw prior to this all their data in the dataset will be set to missing.

### Data collection

To collect our study outcomes and related data (i.e. patient characteristics) the research assistants present at both intervention and existing care facilities will observe each recruited patient’s consultation with the relevant HCPs. They will collect all data via a questionnaire, running on tablet computers, that was developed using RedCap digital survey software and pre-tested internally. Additionally, they will also collect prescription data for patients who are prescribed any medicines, which will be collected from the patient’s paper prescription.

### Data management and quality

All quantitative data will be recorded via mobile digital tablets by the field data collectors, using the RedCap digital survey application. We will use an online data tracker to monitor timely data collection at all the sites. The data will then be exported to a Stata format database for analysis. Overall data quality will be ensured through in-depth training and supervision of data collectors from the research team. The quality of the quantitative data will be maximised by careful design of the data collection tool, including automated checks, and extensive pre-testing.

### Statistical analyses

For all analyses we will base our statistical inferences on the 95% confidence intervals for our estimates of interest. We will use Stata(23) and R(24) statistical software for all data processing and analysis.

#### Primary and secondary outcomes

We will estimate the effect of the intervention at 6- and 12-months after implementation on our primary and secondary outcomes using a standard linear regression model for estimating a difference-in-differences effect(21) (sometimes referred to as a linear probability model when used to analyse binary outcomes, which our secondary outcomes all are). The model will include covariates for study period (either baseline/6-months or baseline/12-months, depending on what study period the effect of interest relates to), treatment group (intervention/existing care) and the interaction between study period and treatment group, with the estimator of the causal effect of the intervention (on the additive scale) given by the interaction coefficient.

To try and reduce confounding bias the model will also contain covariates for patient age and patient age^2^, patient sex (female/male), and date of visit and date of visit^2^ (both measured in terms of the number of days relative to the date of the visit of the first patient in the dataset). Given the clustered (facility-based) sampling of patients we will compute clustered standard errors (at the facility-level) with the HC3 correction for the model coefficients(22). We will compute 95% confidence intervals for our coefficients of interest based on the t-distribution using these standard errors. Aside from patients who do not consent to participate, we do not expect any missing data to occur other than through accidental omission. Therefore, if any missing data are present we will in the first instance carry out complete case analyses as the amount of any missing data should be very minimal.

We will also explore possible effect modification of the intervention effect by facility type (UD vs NGO clinic) and patient sex (female vs male). In both cases we will first compute the intervention effect via the above approach for subgroup separately, and then the difference between those two subgroup-specific intervention effects via difference-in-difference-in-differences or triple difference estimator(25). As these analyses will be underpowered they will be treated strictly as exploratory.

#### Simple app usage

We will estimate the usage of the Simple App by computing the percentage of patients in the intervention group at 6-months and at 12-months who are recorded in the Simple App, along with 95% confidence intervals for these point estimates. We will compute each 95% confidence interval by fitting an intercept-only logistic regression model to the outcome and computing the Wald-based confidence interval on the log-odds scale before transforming it to the probability scale and multiplying by 100, with complex survey design Taylor series linearisation methods used to adjust the confidence interval for the clustered sampling(26).

#### Healthcare provider knowledge

We will estimate the change in HCP knowledge before and after the intervention training programme by computing the mean of the changes in HCP knowledge scores, along with the 95% confidence interval for this point estimate (based on the t-distribution).

### Evaluation of other RE-AIM domains

To evaluate the other RE-AIM domains (other than effectiveness) we will use a mix of qualitative and quantitative data. The quantitative data will either be from the effectiveness study data described above or described separately below. The qualitative data will come from in-depth interviews with patients and stakeholders. These are explained in more detail in each section below.

## Reach

### Quantitative data

We will assess the reach of the intervention in several ways. First, we will use study facility records to look at the total number of individuals aged ≥ 40 (and also disaggregated by sex, age and red card holder status) who would likely benefit each year from the intervention across just the study facilities and (by extrapolating) then also including those facilities where the intervention may be scaled-up to in the future, conditional on our evaluation and future evidence and decisions. Second, more speculatively, we will also look at how many individuals aged ≥ 40 (and also disaggregated by sex, age and red card holder status) are living within the catchment areas of both the study facilities and those facilities where the intervention might be scaled-up to. These individuals would therefore have the potential to benefit from the intervention at some point, should they visit a UD or NGO type PHC facility, conditional on scale-up. We will obtain this data from the census records for catchment area populations available from the DNCC, DSCC and the Civil Surgeon Office. The latest patient census was conducted in 2022 and contains ward-wise population sizes disaggregated by sex in both the city corporations. Third, we will also describe the distribution of key characteristics among patients attending the study facilities, such as sex, age, and red card status.

### Qualitative data

To understand why the intervention may reach some patients and not others we will identify an UD and an NGO clinic with better reach and one of each facility type with lesser reach, based on the observation data collected in 6-months post-intervention. We will then interview a sample of 8 HCPs purposively selected from these facilities. The interviews will explore any differences that they perceive in the types of patients accessing and benefiting from the intervention and whether there are some in the facility catchment area that they believe may not be benefitting from the intervention and their perceptions of why this may be. Interview guides will also explore whether the HCPs report any challenges in delivering the lifestyle counselling to specific groups of patients (e.g. by gender, age, literacy level or socio-economic status).

Additionally, a sample of 14 patients aged ≥ 40 attending the 4 purposively selected intervention facilities will be interviewed after they exit the facilities to identify any differences in their experience while receiving the intervention. Participants will be purposefully sampled based on their age, gender, and whether they are a red-card holder or not, and their NCD status, to understand their experience of screening.

### Adoption

#### Quantitative data

The adoption of the intervention by facilities and by staff within the facilities will be assessed by measuring the number and proportion of facilities invited to participate in the intervention group that accepted or declined, and the number and proportion of HCPs invited for training that accepted and declined. Following training, we will also measure the number and proportion of facilities and HCPs that begin and do not begin implementation, and the number and proportion that completely stop delivering the intervention.

#### Qualitative data

We will purposively select and interview facility managers (in-charges) and HCPs that have not adopted the intervention, or one or more specific components of the intervention, to explore the reasons for the lack of adoption. We will also purposively select and interview facility managers and HCPs that have fully adopted to the intervention to understand their motivations for taking up the intervention.

### Implementation

#### Quantitative data

In the RE-AIM framework the implementation domain focuses on the fidelity and consistency with which the intervention is delivered as intended. Therefore, as our effectiveness outcomes look at whether the different patient management processes that are intended to occur under the intervention are being appropriately carried out or not, our primary and particularly secondary outcomes will also therefore be used to assess this domain. However, whereas the assessment of the effectiveness domain will involve comparing these outcomes between the intervention and existing care groups, for the assessment of the implementation domain we will use these outcomes to describe the level of fidelity to the intervention components occurring within the intervention group at the patient- and facility-level, and how this fidelity varies across all facilities and between facility type at 6-months post-intervention.

#### Qualitative data Health care providers

During the HCP training programme, a researcher will be present to observe the process of training and qualitatively document any observed facilitators and barriers during the sessions. We will also aim to interview 8 HCPs who receive the intervention training to understand their perceptions regarding the training programme.

We will also identify a UD and an NGO clinic showing better adoption of the intervention, and another UD and an NGO clinic showing poorer adoption, based on the outcome data collected at 6-months post-intervention. Within these facilities, qualitative observations will be conducted by a senior qualitative researcher to observe the implementation of the intervention. This will be done to identify factors which may be affecting the implementation of the intervention within the facility, such as lack of privacy, noise, lack of electricity, lack of washrooms, and shortages of medications or diagnostic equipment.

Within these same facilities we will identify one HCP and one facility manager from each facility and interview them about their experiences of implementing the intervention, focusing on the facilitators and barriers to implementation of all components of the intervention that they report. Both managers and HCPs will also be asked to share their perceptions of any unintended consequences (positive or negative) of including the intervention within their routine practice in the facility (e.g. lack of time for other activities, better relations with patients or referral institutions).

#### Patients

As described in the *Reach* section above, in the patient exit interviews we will also ask those 14 patients about their experiences of the intervention to understand which elements of the intervention they remember receiving and any reflections of their interaction with the HCP and the facility. We will also ask those patients how confident they feel to be to adhere to the advice given to help them manage their diabetes and/or hypertension and any challenges they think they will experience. We will also ask them about any costs they have incurred during their visit or in buying medications.

#### Policy makers

We will interview policy makers, including government officials and development partners, about their opinions regarding the intervention. Specifically, we will seek to understand how they found the intervention co-creation process, why they wanted to base it on the National Protocol and Simple App, how it has been working in rural facilities, the challenges we have found it may be facing in urban facilities, and their views on how we might overcome or mitigate those challenges to allow scale-up and ensure sustainability.

### Maintenance

#### Quantitative data

To assess maintenance of the intervention we will also make use of the effectiveness outcomes. However, similar to our use of them to assess the implementation domain, we will use these outcomes to describe the level of fidelity to the intervention components occurring within the intervention group at the patient and facility-level, and how this fidelity varies across all facilities and between facility type, but now at 12-months post-intervention. To further assess maintenance, we will also evaluate the longer-term effectiveness of the intervention via our 12-months post-intervention effectiveness analyses.

#### Qualitative data

We will again interview the HCPs sampled for the implementation domain qualitative interviews but now after the implementation period, to understand reasons for continuation or discontinuation of the intervention and the separate components of the intervention. Simultaneously, we will also interview HCPs in existing care facilities to understand the reasons for any changes, or lack of changes, in their NCD management practices and data recording. This will help us to assess whether there has been any contamination between the intervention and existing care groups.

### Data analyses for other RE-AIM domains’ data

#### Quantitative data

For the reach and adoption domain we will estimate simple summary statistics (counts and percentages) appropriate for the planned measures, plus 95% confidence intervals for the percentages as appropriate. For the implementation and maintenance domain descriptive analyses we will summarise all outcomes in terms of percentages and their 95% confidence intervals, and for just the maintenance domain we will estimate intervention effects at 12-months as described in the *Statistical analyses* section for the effectiveness domain.

#### Qualitative data

We will transcribe all interviews verbatim in Bengali and then translate those transcriptions into English for members of the research team who do not speak Bengali. This will be done for quality assurance and transparency of data among Bengali-speaking and non-Bengali-speaking members of the research team. We will use the Framework Approach (27) with an a-priori framework structured according to the RE-AIM domains. We will generate codes inductively from our data and group them within each of these broad domains. The coding will be done by three trained qualitative researchers who will start by familiarising themselves with each transcript. Two of the researchers will initially code the transcripts independently. The third coder will then assist in resolving any differences in coding. They will then all discuss the codes and agree how they can best be grouped within the framework of RE-AIM domains. Once the codes are finalised, they will then apply the framework to all the qualitative data collected from both HCPs, managers and patients. We will use NVivo(28) Software to conduct qualitative data analysis.

They will then develop narrative summaries under each of the domains of the RE-AIM framework, which will highlight differences in findings between facility types, and between patients of different genders, socio-economic backgrounds, and NCD statuses (those with hypertension and/or diabetes). This will inform the respective RE-AIM domains from an equity perspective. To integrate our quantitative and qualitative findings we will develop a meta-inference table using the RE-AIM domains, which will highlight similarities and differences in findings between the different types of data and/or different participant groups (HCPs, managers and patients) (29).

## Qualitative data management and quality

(For details on quantitative data management and quality see *Data management and quality* section above.) We will collect all qualitative interview data using digital audio recorders, with the data transferred to the password-protected computer of the qualitative researchers. The recording will be deleted from the recorder once the verbatim transcript is produced. Overall data quality will again be ensured through in-depth training and supervision of data collectors from the research team, focusing on good questioning and probing techniques for the qualitative data collectors. Detailed feedback will also be provided on the qualitative transcripts and analyses by senior researchers.

## DISCUSSION

In Bangladesh, the prevalence of NCD risk factors, hypertension and diabetes are higher among those living in urban areas compared to those living in rural areas (14,15). The burden of this eventually falls on Bangladesh’s pluralistic urban PHC system(6). Although there are many different types of PHC facility in urban areas, in this study we are focusing on those (the UDs and NGO clinics) which offer services and medications at no or minimum cost to patients. The aim of the planned intervention is to strengthen the ability of these UDs and NGO clinics to manage the rising tide of hypertension and diabetes, and reduce patients’ risk of other cardiovascular diseases, through timely management of patients either at risk of developing these conditions or those found or known to have these conditions, by systematic and effective screening, diagnosis, lifestyle counselling, use of appropriate medication, and effective follow-up and referral processes. The longer-term goal is for this evidence, if positive, to justify the government to support scale-up of the intervention across all such facilities in urban Bangladesh.

## ETHICS AND DISSEMINATION

Ethics approval has been received from the Research Governance Committee at the University of Leeds, UK (MREC 21-008) and from the Bangladesh Medical Research Council (BMRCAIREC/20 I 9 -2022 / 485). We will use a variety of channels to share our findings with policy makers, service providers, academicians, and relevant stakeholders.

## Supporting information

Supplemental material (Theory of Change)

## Authors’ Contributors

DB, RH, JH and HE conceived the study and wrote the first and all subsequent drafts. TE, BE, MD KI, RC, MA, USA and NBJ, made comments and suggestions on the draft paper. All authors participated in manuscript revisions, edits and read and approved the final manuscript.

## Funding Statement

This study is funded by a grant received from UK Aid from the UK Government, Grant 301132; however, the views expressed do not necessarily reflect the UK government’s official policies’

## Competing interests

None declared.

## Patient and public involvement

Patients and/or the public will be involved in the design and intervention co-creation, reporting, and dissemination plans of this research.

## Data availability statement

Data are available upon reasonable request.

## Notes

### Competing Interest Statement

The authors have declared no competing interest.

### Author Declarations

Ethics approval has been received from the Research Governance Committee at the University of Leeds, UK (MREC 21-008) and from the Bangladesh Medical Research Council (BMRCAIREC/20 I 9 -2022 / 485).

