## Supplemental material (Theory of Change) for "Understanding The Impact, Reach and Implementation of a Health Systems Intervention to Improve Diabetes and Hypertension Care in Pluralistic Urban Public Primary Care in Bangladesh: A Study Protocol"

Supplementary Material: 1 (Theory of Change)

Theory of Change

| INPUTS | ACTIVITIES | OUTPUTS | OUTCOMES | IMPACT |
| --- | --- | --- | --- | --- |
| - PEN adapted national protocol for management of hypertension and diabetes at primary health care centres - Simple App | - Signing MoU between ARK and NCDC, MoHFW for implementation of protocol at urban primary health care centres - Signing MoU between ARK and National Heart Foundation for use of Simple app at urban primary health care centres - Creating steering committee with members from MoHFW, and MoLGRD&C - Communicating and coordinating with: - Representatives of NCDC, MoHFW who are involved in the development of materials, training, and implementation of PEN adapted National Protocol - Representatives of Dhaka Civil Surgeon Office who are responsible for Government Outdoor Dispensaries - Representatives of Urban Primary Health Care Management Unit of MoLGRD&C who are responsible for NGO clinics under the Urban Primary Health Care Service Delivery Project - Representatives of World Health Organization (WHO) Country office for PEN - Representatives of RESOLVE TO SAVE LIVES and National Heart Foundation who are responsible for implementation of Simple application at rural primary health care centres - Facility managers and health care workforce of Government Outdoor Dispensaries and NGO clinics - Discussing intervention implementation pathways - Accessing protocols, training and lifestyle modification materials, NCD registers, and Simple application - Preparing for training: - Communicating with potential trainers - Finalizing dates, venues, and duration, - Choosing facilities for training - Choosing health workforce from selected facilities for training - Preparing training agenda and slides - Procuring logistics such as tabs for simple app, and protocols | Urban primary Health Care Providers trained on protocol | Urban PHC health workforce has updated knowledge about NCD management | Capacity of heath workforce to manage NCDs strengthened |
|  |  | Improved skills of healthcare workers in:   - Identifying patients at risk of having CVD within 10 years - Identifying patients at risk of being hypertensive - Identifying patients at risk of being diabetic | Increased screening for:   - Early detection of persons aged ≥ 40 at NCD risk - Early diagnosis of NCD patients aged ≥ 40 | Prevention of:   - NCDs among those at risk, thereby preventing the patient in having to bear additional expense due to NCD drugs - Complications among NCD patients, especially those who are unaware about their NCD status |
|  |  | Updated skills of healthcare workers in identifying:   - Hypertensive patients - Diabetic patients |  |  |
|  |  | Improved skills of healthcare workers in counselling those at risk of NCDs and NCD patients about lifestyle modification | Increased lifestyle modification counselling of:   - persons aged ≥ 40 at risk of developing NCDs, - NCD patients aged ≥ 40 |  |
|  |  | Updated (improved) knowledge of:   - Doctors in prescribing NCD medications - Other HCP on advising patients for follow up - Doctors in referring patients | Patients receiving updated:   - Treatment - Advice regarding follow up - Advice regarding referral |  |
|  |  | Urban primary health care centres are provided with:   - Tabs with Simple app | Following data regarding NCD patients are recorded:   - Socio demographic characteristics - Behaviour risk factors of NCD patients - Height, weight, Blood pressure and Blood glucose levels | Transfer of data from urban primary health care centres to the national health dashboard can help policy makers stay informed about:   - NCD status among urban dwellers, segregated by sex, and age - Prevalence of behavioural risk factors among NCD patients - BMI, blood pressure and blood glucose trends among NCD patients |
|  |  |  | Medicines prescribed to NCD patients recorded | NCDC programme can have an idea about the quantity of NCD medications required at urban primary health care centres |
|  |  |  | Follow up advice given to the NCD patient recorded | Loss to follow up of patients can be prevented |
